# Determinants of prices of commercial electric breast pumps sold in Germany: a cross-sectional analysis

**DOI:** 10.64898/2026.11.13.26367926

**Authors:** Jelica Gencel-Augusto, Elochukwu Okanmelu, Frances Strobl, Monika Ziebart, Melissa A. Theurich

## Abstract

In Germany, statutory health insurance covers the rental or purchase of breast pumps when a medical indication is present for the mother or infant, and a physician has issued a prescription. However, parents must pay out of pocket for commercially available breast pumps for returning to work or personal circumstances. High indirect costs for continuation of breastfeeding and direct costs for breast pumps and related equipment may pose substantial barriers to breastfeeding families, an issue relevant for health equity. To our knowledge, no studies have assessed the direct financial costs associated with the purchase of commercially available breast pumps. In this cross-sectional study, we examined digital marketplaces in Germany for commercial electric breast pumps between March 2025 and May 2026, across major online platforms. Our objective was to explore pump features that predict price. We identified 282 commercial electric pump models from 108 brands. Basic information was unavailable for half of surveyed breast pump brands. The median price for surveyed commercial electric pumps was €72.70 (IQR: €82.70), with costs ranging widely from €19.99 to €659.57. Prices varied by marketplace, manufacturer location, accessories, pump system type, power source, battery capacity, flange sizes, availability of instruction manuals, and “single user” claims; predicting significantly higher costs. Half of pumps surveyed offered only one flange size, signaling additional expenses for mothers needing alternative sizes. These findings highlight important considerations for maternal health and consumer safety. Health technology assessments and a review of current health policy on reimbursement for commercial electric breast pumps is needed.

**Key messages:**

- To our knowledge, there have been no published empirical studies on direct costs for commercial electric breast pumps
- Basic information was unavailable for half of the surveyed breast pump brands on digital marketplaces in Germany
- The median cost for a commercial electric breast pump in our survey was €72,70 (IQR: €82,70), with a very wide price range (€19,99 - €659,57)
- Prices varied significantly by marketplace, manufacturer location, accessories, pump system type, power source, battery capacity, number of flange sizes, availability of instruction manuals, and “single user” claims, which all positively correlated with prices.
- There is a sparsity of flange sizes sold outside of the mid-range (24 mm), signaling ancillary costs for flanges with appropriate fit
- Health technology assessments for commercial electric breast pumps and a review of health policy related to breast pump reimbursement in Germany are warranted

## 1. Introduction

Healthcare systems indisputably save money on healthcare costs when families breastfeed, through both the prevention and lessening of the burden of diseases and associated healthcare expenditures (1–5). Yet scientific literature has paid far less attention to estimating the direct costs of provision of expressed breast milk to families (6). In recent years, studies have demonstrated that an increasing number of lactating women report expression of breast milk using breast pumps (7–10). In Germany, prescriptions for breast pumps have steadily increased over the past decade (11). This epidemiological data aligns with commercial market data estimating the German breast pump market at approximately $107.6 million, with projected growth to $170.5 million by 2030 (12). Despite their apparent widespread use, limited empirical research has explored direct costs associated with human milk expression for families. To estimate direct costs, economic studies on breast pumps and related infant feeding equipment are needed.

Broadly speaking, there are two main categories of breast pumps. Multi user, medical grade electric pumps are generally larger, heavier, and equipped with more powerful motors (13). These are typically stationary medical devices that must be plugged into a wall outlet, lack a battery, and are designed for use by multiple individuals, like in healthcare settings (13, 14). In contrast, commercially available electric breast pumps are normally intended for only one user, also known as ‘personal-use’ pumps, are smaller, lighter, and are usually portable with rechargeable batteries (15). Commercially available breast pumps are also less expensive compared to multi-user pumps. These pumps are typically available for purchase in drug stores, pharmacies or online marketplaces (16).

The cost of commercial breast pumps are an important consideration since statutory health insurance, which covers around 90% of the German population (17), only reimburses breast pump expenses if there is a medical indication in the mother or infant (18). This means that pumps purchased for reasons such as returning to work or school, or lifestyle preferences are not reimbursed (18). As a result, commercial breast pumps electively acquired must be paid by parents out of pocket (18).

To our knowledge, no empirical studies have examined the direct costs of commercially available breast pumps in European contexts. However, this information is essential to understand the potential impact of breast pump costs on parental decisions for the continuation of breastfeeding. It is also important for healthcare professionals supporting parents in informed decision making on infant feeding. The objective of this study is to explore commercially available electric breast pumps sold in Germany, describing their characteristics and costs. Secondary objectives are to analyze the pump characteristics that predict cost and explore the flange sizes sold together with commercial electric breast pumps.

## 2. Methods

A cross-sectional study was conducted to explore and describe the range of commercially available breast pumps sold on digital marketplaces in Germany. A cost analysis was conducted to examine the relationship between cost and breast pump characteristics. This study is reported based on a modified version of the ‘Consolidated Health Economic Evaluation Reporting Standards’ (CHEERS II) statement (19). Items 4, 13, 14, 16, 17, 19, 21, 25 were not included since they were not applicable to our study.

### 2.1 Protocol and registration

We publicly registered the protocol for this study with the Open Science Framework (OSF) before conducting data analysis (20).

### 2.2 Study population, setting and location

Although no participants were enrolled in this study, its findings are directly relevant to women of reproductive age living in Germany, from varying socioeconomic status (SES), who plan to use a breast pump. The setting of this study is German digital marketplaces. We expected a wider variety of commercially available electric breast pumps to be sold online, compared to brick and mortar stores.

### 2.3 Inclusion and exclusion criteria

Commercially available breast pumps with an electric motor were included. Medical-grade pumps and manual pumps were excluded.

### 2.4 Outcome variable

The primary outcome was price. A cross-sectional approach was used due to the volatility of price data, effects of discounts, and rapid product turnover on digital marketplaces. We recorded prices from March to September 2025 in Euros (EUR). We recorded the Recommended Retail Price (RRP) in EUR and discount rate (%) where applicable. Discount rates were applied to final analysis to reflect real-time price of pumps.

### 2.5 Predictor variables

We extracted predetermined variables on breast pumps, including the brand name, brand headquarter country, country of manufacturer, and availability of the Conformité Européenne (CE) safety certificate.

Technical characteristics of pumps were recorded including: the pumping system configuration (open/closed), wearable versus non-wearable design, single- or double-sided, pump size (in m^3^), battery type (rechargeable or not), battery capacity (mAh), maximum noise level (dB), weight (grams), minimum and maximum vacuum pressure (mmHg), number of milk expression modes, number of suction intensity levels, volume of milk collection containers (in mL), availability of flanges, and flange number and sizes.

We recorded whether the breast pump was intended for a single user or multiple users, and whether resale was prohibited. Marketing claims, such as ‘hands-free’ and ‘BPA-free’ were extracted.

### 2.6 Search strategy

#### Data sources

We searched the following websites: amazon.de, otto.de, kaufland.de, mediamarkt.de/saturn.de, conrad.de, baby-walz.de, babyone.de, babyshoppen.de, babymarkt.de, rossmann.de, mueller.de, dm.de, medpex.de, shop-apotheke.com, docmorris.de, marktkauf.de, netto-online.de, lidl.de, aldi-nord.de/aldi-sued.de for pumps available online in Germany. We identified brand websites. Where available, we then also searched each brand website.

We used the search term ‘electric breast pump’ (e.g., German language: ‘*elektrische Milchpumpe’*). Search results frequently included products other than electric breast pumps (e.g., accessories, manual breast pumps, and replacement parts), which were excluded from the study.

#### Data collection and extraction

A master list was developed in Microsoft Excel comprising all eligible breast pump listings identified across all websites. Breast pump data were taken directly from the brand’s website or marketplace. Where information was inconsistent between the brand website and the marketplace, we recorded both but used information from the brand website for the analysis. Where information was missing, we searched the instruction manual. Data was extracted by one reviewer (JGA) and was checked by two others (MT, EO). Extracted data was in German language but translated into English for the purpose of writing this manuscript.

### 2.7 Data analysis

Data analysis was performed using R-programming version 4.3.3. Distribution of data was checked using density plots, QQ plots and boxplots. Price exhibited right-skewness and heteroscedasticity. Descriptive statistics (median and interquartile range) were used for continuous variables, while frequency and percentage distribution were used for categorical variables. Overall differences in price distributions were evaluated using Mann-Whitney U (Wilcoxon rank-sum test) and the Kruskal–Wallis tests. Pairwise associations were assessed using Pearson’s chi-squared test. Holm-adjusted p-values were used in cases for multiple comparisons (21). A Spearman rank correlation was conducted to assess the relationship between continuous variables and price. Multivariate linear regressions were fit to assess predictors of price. Regression analyses modeled a natural logarithmic of price (log-price: %Δ=(e^β^-1) ×100) as the dependent variable. A backward selection procedure based on Akaike Information Criterion (AIC) was applied for model selection. Regression coefficients were subsequently interpreted as percentage changes in price using exponentiated estimates (22).

### 2.8 Sensitivity analysis

Given the paucity of studies on this topic, we conducted a sensitivity analysis for predictor variables for the pumps with missing values. Missing data in regression analysis was handled through multiple imputations using predictive mean matching. Imputation was restricted to independent variables with less than 30% missingness (23). Predictor variables with more than 30% missingness were excluded from the main analysis. Missingness is reported in **Supplementary Figure 1: Overview of Missing and Observed Data for Commercial Pumps on German Digital Marketplaces.**

### 2.9 Photographs

We chose a very small, purposive selection of commercially available electric breast pumps to photograph. We based the selection on innovations in the breast pump design (multi-purpose milk collection vessels, tubing, handsfree design, remote control, touchscreen/buttons, control through a mobile device, etc.).

## 3 Results

### 3.1 Included commercial electric breast pumps

In all, 282 commercially available electric breast pumps were identified from 108 brands. A total of 73 websites were searched, including 54 brand websites and 19 online marketplaces. Digital marketplaces include electronic stores, pharmacies, supermarkets, infant or maternity online stores, and general multi-vendor marketplaces. A flowchart of the selection process is outlined in **Supplementary Figure 2: Flowchart of selection process for commercial electric breast pumps on digital marketplaces.**

### 3.2 Brands and manufacturers

Of the 108 identified breast pump brands, company headquarter addresses were provided by 66 brands (61%), but only half (*n=*54, 50%) had an official company website. Of the 66 brands with a physical address, just over half (*n=*38, 58%) were based in Europe, commonly in Poland (*n*=9, 14%), with only two brands (3%) headquartered in Germany.

Manufacturing countries were identified for just over half of the surveyed brands (*n=*61, 56%), with China being the most prevalent country of manufacturing (*n=*51 brands). A comprehensive list of surveyed breast pumps brands, with headquarters and manufacturer information are listed in **Supplementary Table 1 Breast pump brand headquarters and manufacturing country information.**

### 3.3 Characteristics and technological features of commercial electric breast pumps

The proportion of single-sided vs double-sided pumps was similar, with approximately half (*n*=144, 51.1%) being single-sided. There were 123 (43.6%) wearable pumps. Breast pumps were classified as ‘wearable’ if they were designed to be worn entirely inside the bra, with an integrated compartment to collect expressed breast milk and without external tubing. ‘Hands-free’ claims were common (*n*=148, 52.5%). Pumps had milk collection containers holding up to a median volume of 180 ml (IQR: 30) of expressed breast milk. The median weight of breast pumps was 275g (IQR: 129.9). **See Table 1**: Descriptive characteristics of breast pump brands with at least four breast pumps

**Table 1:**
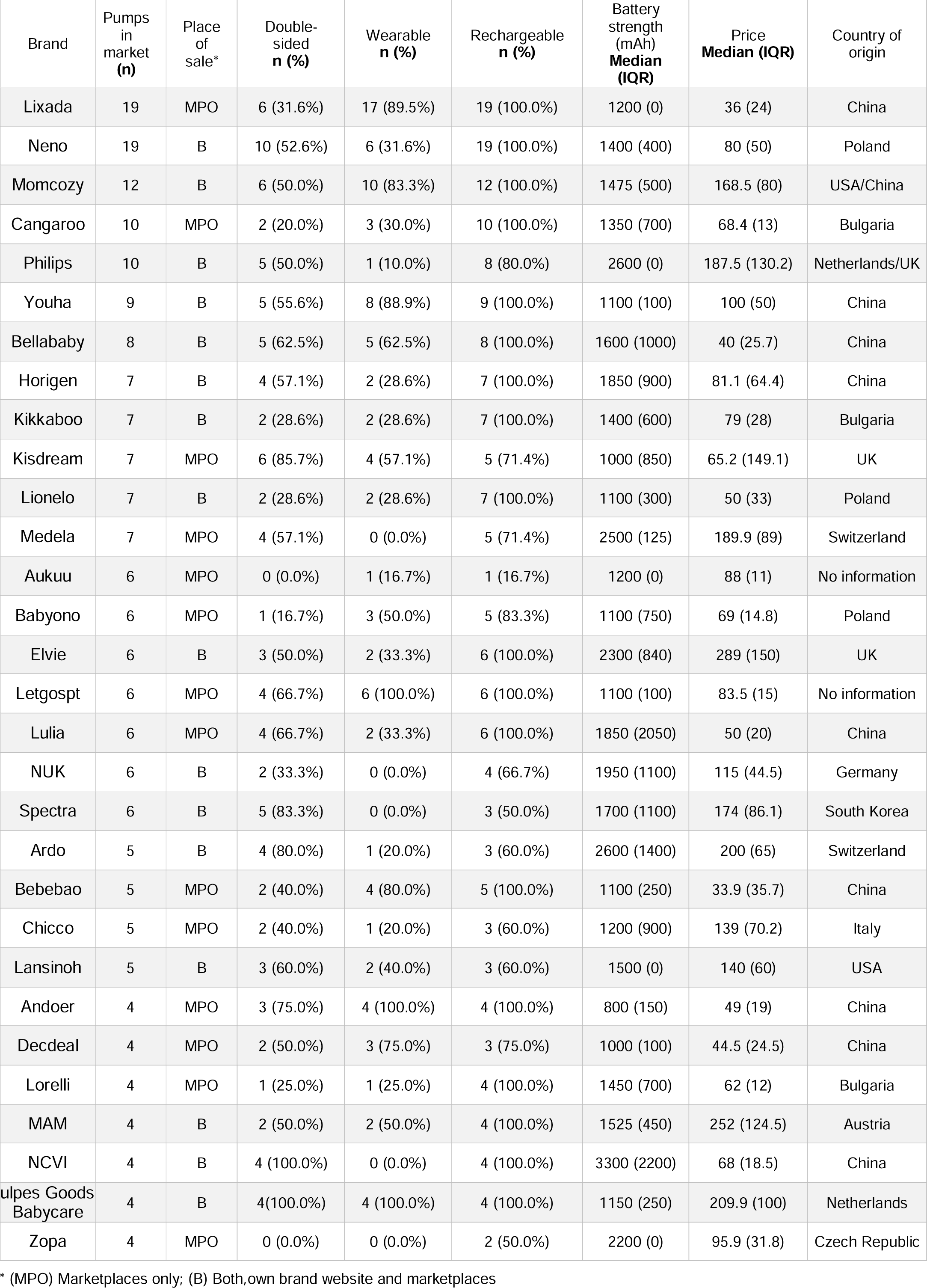
Descriptive characteristics of breast pump brands with at least four breast pumps.

| Brand | Pumps in market **(n)** | Place of sale* | Double-sided **n (%)** | Wearable **n (%)** | Rechargeable **n (%)** | Battery strength (mAh) **Median (IQR)** | Price **Median (IQR)** | Country of origin |
|---|---|---|---|---|---|---|---|---|
| Lixada | 19 | MPO | 6 (31.6%) | 17 (89.5%) | 19 (100.0%) | 1200 (0) | 36 (24) | China |
| Neno | 19 | B | 10 (52.6%) | 6 (31.6%) | 19 (100.0%) | 1400 (400) | 80 (50) | Poland |
| Momcozy | 12 | B | 6 (50.0%) | 10 (83.3%) | 12 (100.0%) | 1475 (500) | 168.5 (80) | USA/China |
| Cangaroo | 10 | MPO | 2 (20.0%) | 3 (30.0%) | 10 (100.0%) | 1350 (700) | 68.4 (13) | Bulgaria |
| Philips | 10 | B | 5 (50.0%) | 1 (10.0%) | 8 (80.0%) | 2600 (0) | 187.5 (130.2) | Netherlands/UK |
| Youha | 9 | B | 5 (55.6%) | 8 (88.9%) | 9 (100.0%) | 1100 (100) | 100 (50) | China |
| Bellababy | 8 | B | 5 (62.5%) | 5 (62.5%) | 8 (100.0%) | 1600 (1000) | 40 (25.7) | China |
| Horigen | 7 | B | 4 (57.1%) | 2 (28.6%) | 7 (100.0%) | 1850 (900) | 81.1 (64.4) | China |
| Kikkaboo | 7 | B | 2 (28.6%) | 2 (28.6%) | 7 (100.0%) | 1400 (600) | 79 (28) | Bulgaria |
| Kisdream | 7 | MPO | 6 (85.7%) | 4 (57.1%) | 5 (71.4%) | 1000 (850) | 65.2 (149.1) | UK |
| Lionelo | 7 | B | 2 (28.6%) | 2 (28.6%) | 7 (100.0%) | 1100 (300) | 50 (33) | Poland |
| Medela | 7 | MPO | 4 (57.1%) | 0 (0.0%) | 5 (71.4%) | 2500 (125) | 189.9 (89) | Switzerland |
| Aukuu | 6 | MPO | 0 (0.0%) | 1 (16.7%) | 1 (16.7%) | 1200 (0) | 88 (11) | No information |
| Babyono | 6 | MPO | 1 (16.7%) | 3 (50.0%) | 5 (83.3%) | 1100 (750) | 69 (14.8) | Poland |
| Elvie | 6 | B | 3 (50.0%) | 2 (33.3%) | 6 (100.0%) | 2300 (840) | 289 (150) | UK |
| Letgospt | 6 | MPO | 4 (66.7%) | 6 (100.0%) | 6 (100.0%) | 1100 (100) | 83.5 (15) | No information |
| Lulia | 6 | MPO | 4 (66.7%) | 2 (33.3%) | 6 (100.0%) | 1850 (2050) | 50 (20) | China |
| NUK | 6 | B | 2 (33.3%) | 0 (0.0%) | 4 (66.7%) | 1950 (1100) | 115 (44.5) | Germany |
| Spectra | 6 | B | 5 (83.3%) | 0 (0.0%) | 3 (50.0%) | 1700 (1100) | 174 (86.1) | South Korea |
| Ardo | 5 | B | 4 (80.0%) | 1 (20.0%) | 3 (60.0%) | 2600 (1400) | 200 (65) | Switzerland |
| Bebebao | 5 | MPO | 2 (40.0%) | 4 (80.0%) | 5 (100.0%) | 1100 (250) | 33.9 (35.7) | China |
| Chicco | 5 | MPO | 2 (40.0%) | 1 (20.0%) | 3 (60.0%) | 1200 (900) | 139 (70.2) | Italy |
| Lansinoh | 5 | B | 3 (60.0%) | 2 (40.0%) | 3 (60.0%) | 1500 (0) | 140 (60) | USA |
| Andoer | 4 | MPO | 3 (75.0%) | 4 (100.0%) | 4 (100.0%) | 800 (150) | 49 (19) | China |
| Decdeal | 4 | MPO | 2 (50.0%) | 3 (75.0%) | 3 (75.0%) | 1000 (100) | 44.5 (24.5) | China |
| Lorelli | 4 | MPO | 1 (25.0%) | 1 (25.0%) | 4 (100.0%) | 1450 (700) | 62 (12) | Bulgaria |
| MAM | 4 | B | 2 (50.0%) | 2 (50.0%) | 4 (100.0%) | 1525 (450) | 252 (124.5) | Austria |
| NCVI | 4 | B | 4 (100.0%) | 0 (0.0%) | 4 (100.0%) | 3300 (2200) | 68 (18.5) | China |
| [illegible]ulpes Goods Babycare | 4 | B | 4(100.0%) | 4 (100.0%) | 4 (100.0%) | 1150 (250) | 209.9 (100) | Netherlands |
| Zopa | 4 | MPO | 0 (0.0%) | 0 (0.0%) | 2 (50.0%) | 2200 (0) | 95.9 (31.8) | Czech Republic |

\* (MPO) Marketplaces only; (B) Both,own brand website and marketplaces

**Table 2:** Commercial breast pump characteristics predicting price.

| Predictor | Estimate | CI | Percent change | Percent change CI | p-value |
|---|---|---|---|---|---|
| Intercept | 3.901 | 3.413 to 4.389 | 4846.1 | 2937% to 7955.3% | 0 |
| Collection volume (mL) | -0.002 | -0.004 to 0 | -0.2 | -0.4% to 0% | 0.05 |
| Marketplace (vs brand website) | -0.2 | -0.338 to -0.062 | -18.1 | -28.7% to -6% | 0.00483 |
| Marketplace & brand website (vs brand website) | -0.352 | -0.603 to -0.102 | -29.7 | -45.3% to -9.7% | 0.00603 |
| Battery capacity (mAh) | 0 | 0 to 0 | 0 | 0% to 0% | 0.00145 |
| Additional accessories | 0.131 | -0.057 to 0.318 | 14 | -5.5% to 37.5% | 0.17 |
| Wearable design | 0.209 | -0.01 to 0.427 | 23.2 | -1% to 53.2% | 0.061 |
| Double pump | 0.438 | 0.319 to 0.556 | 54.9 | 37.5% to 74.4% | 0 |
| Hands-free claim | 0.255 | 0.056 to 0.453 | 29 | 5.8% to 57.4% | 0.0121 |
| Single-user claim | 0.086 | -0.103 to 0.275 | 9 | -9.8% to 31.7% | 0.37 |
| Resale prohibited | 0.195 | 0.015 to 0.374 | 21.5 | 1.6% to 45.4% | 0.0334 |
| High operating modes | -0.238 | -0.385 to -0.091 | -21.2 | -31.9% to -8.7% | 0.00161 |
| Instruction manual | 0.248 | 0.052 to 0.444 | 28.2 | 5.3% to 55.9% | 0.0135 |
| BPA-free claim | -0.043 | -0.174 to 0.088 | -4.2 | -16% to 9.2% | 0.521 |
| Asia (vs Europe) | -0.302 | -0.469 to -0.134 | -26 | -37.4% to -12.5% | 0.000512 |
| North America (vs Europe) | 0.373 | -0.009 to 0.755 | 45.3 | -0.9% to 112.8% | 0.0553 |
| No information (vs Europe) | -0.45 | -0.735 to -0.164 | -36.2 | -52.1% to -15.2% | 0.00218 |
| 2 intensity levels (vs 1) | -0.116 | -0.337 to 0.105 | -11 | -28.6% to 11% | 0.299 |
| 3 intensity levels (vs 1) | 0.042 | -0.263 to 0.346 | 4.2 | -23.1% to 41.3% | 0.788 |
| 4 intensity levels (vs 1) | -0.511 | -0.917 to -0.105 | -40 | -60% to -9.9% | 0.0139 |
| Closed + anti-backflow (vs anti-backflow) | 0.219 | 0.017 to 0.42 | 24.4 | 1.7% to 52.3% | 0.034 |
| Closed system (vs anti-backflow) | 0.048 | -0.129 to 0.225 | 4.9 | -12.1% to 25.3% | 0.595 |
| No claim (vs anti-backflow) | 0.078 | -0.092 to 0.248 | 8.1 | -8.8% to 28.1% | 0.368 |
| Other flow-control claims (vs anti-backflow) | 0.214 | -0.282 to 0.709 | 23.8 | -24.5% to 103.1% | 0.396 |
| Number of flange sizes | 0.011 | -0.055 to 0.078 | 1.2 | -5.3% to 8.1% | 0.732 |

Photographs of a purposive sample of commercial electric breast pumps are depicted in **Figure 1**. Some models offered ‘low noise’ operation modes, touchscreens, memory functions and smart remote control via apps. Additionally, certain pumps included adapters for feeding bottles or breastmilk storage bags. Other pumps were advertised as waterproof or with lightweight configuration.

**Figure 1:**
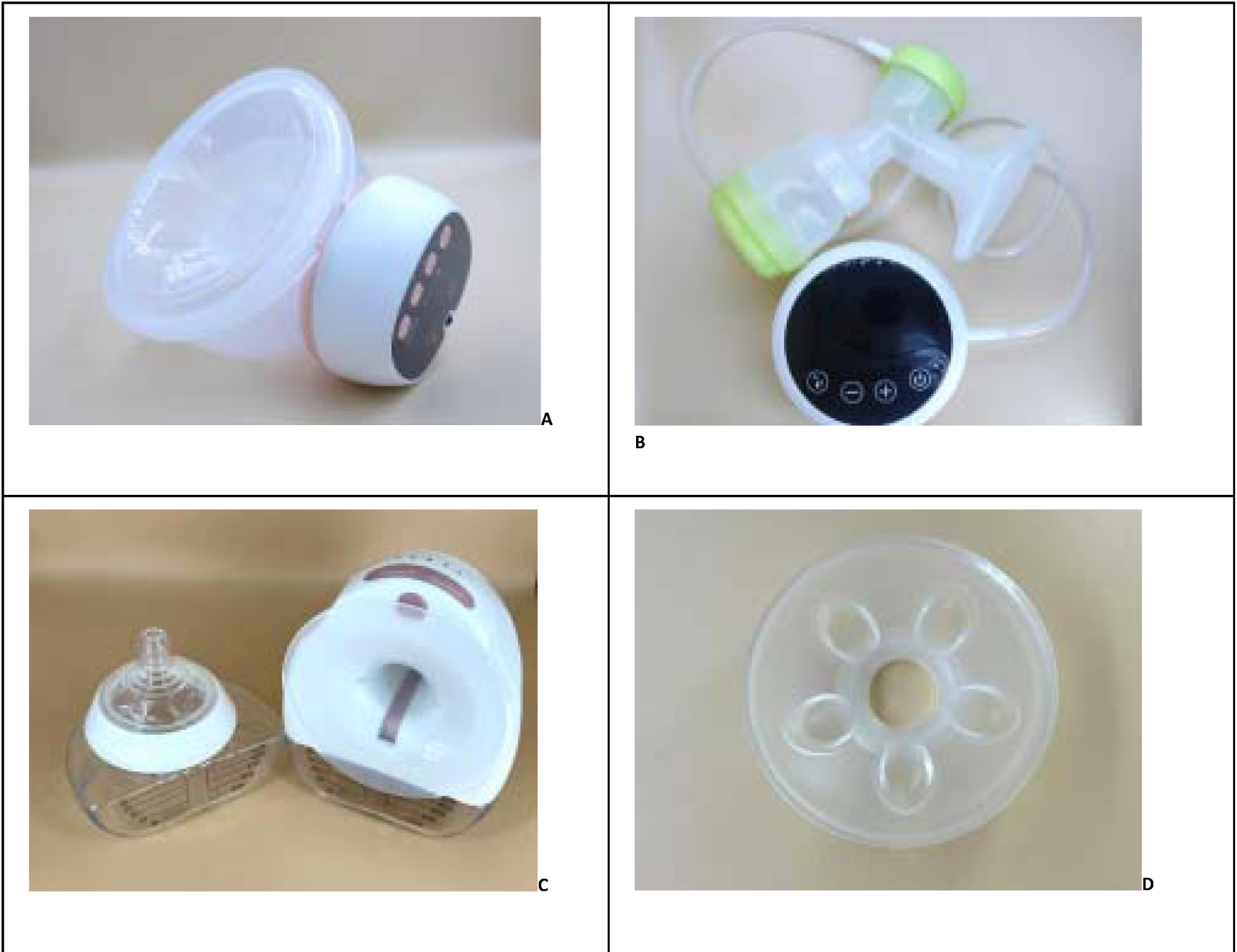
Photographs of commercial electric breast pumps and a ‘universal’ flange. A) One-sided, wearable electric breast pump with display, silicone cushion and plastic cover B) One-sided, electric breast pump with a plastic collection kit and custom-fitting plastic flange C) One-sided, wearable electric breast pump, with a uniform-sized flange, glass milk collection kit and compatible feeding teat D) Uniform-sized silicon cushion

#### Noise and battery life

Noise volume was available for approximately half of pumps (*n*=150, 53.2%). The median noise volume was 48 dB (IQR: 11dB). Most pumps (*n*=241, 85.5%) used integrated lithium rechargeable batteries to function. Battery capacity was reported for 217 of these pumps, with a median capacity of 1300 milliamp-hours (mAh) (IQR: 700 mAh). The median operating time for rechargeable pumps was 120 minutes (IQR: 50). This value was reported for 165 of the 241 rechargeable pumps.

#### Vacuum and cycling

Breast pumps reported a median minimum applied vacuum of 45mmHg (IQR: 25mmHg; *n*=104, 36.9%), and a median maximum of 285mmHg (IQR: 38); (*n*=170, 60.3%). Most breast pumps (*n*=278; 98.6%) mentioned holding multiple ‘Modes of milk expression’ with a median of 3 modes (IQR: 1), and a median of 9 (IQR: 1) adjustable levels of intensity. Some of the modes of milk expression included: *massage, stimulation, pumping, silent pumping, power pumping, deep pumping, expression, suction, deeper suction, deep extraction, sucking, milk drawing, recovery, prolactin mode, dual frequency, automatic, bionic*.

#### Related pumping equipment

Most of the pumps (*n*=252 89.4%) had additional equipment shipped with the breast pump. Apart from bottle accessories (*n*=120, 43%) such as teats, and bottle caps, the most common extra items included with the pump were travel bags for carrying the breast pump (*n*=79, 28%), replacement valves (*n*=78, 28%), bra adjustment buckles (*n*=74, 26%), and breast milk storage bags (*n*=52, 18%). Other less common accessories were flange size rulers (9%), nursing pads (7%), adapters to express milk directly into breast milk bags (4%), brushes to clean pump parts (2%), and remote controls (2%).

#### Safety and hygiene

‘Single-user’ use was specified in just over half of the pumps’ instructions (*n*=149, 52.8%). Although marketed for personal use, the instructions for 5 pumps (1.8%) mentioned that except for the pumping kits, the pump itself could be used by multiple users. In 45 pumps (16%) resale was expressly prohibited, whereas no information on this could be found for the remaining pumps.

Over half of the breast pumps surveyed (*n*=169, 59.9%) provided information on the type of pumping system (open versus closed). The most common terminology used to refer to a pump having a barrier between the pumped breast milk and the internal pump parts was ‘closed system’ (*n*=55, 19.5%) and ‘anti-backflow/anti-reflux system’ (*n*=78, 27.7%). A further 32 pumps (11.3%) claimed both ‘closed’ and ‘anti-backflow/anti-reflux’ systems. No pumps claimed to have an ‘open system’.

Only 172 pumps (61%) displayed the European Union (EU) Conformité Européenne, known as the ‘CE mark. Claims that breast pumps were free from bisphenol A-free (BPA) were reported for most pumps (*n*=211, 75%).

### 3.4 Flange characteristics

A total of 271 (96.1%) breast pumps had data on the total number of flanges, also known as ‘breast shields,’ included with the pump. Half of all pumps (*n*= 155, 55.0%) offered a single flange, followed by two flanges (*n*=51, 18.1%). The median number of flanges provided per pump was 1 (IQR 1-3; range 1-7). Information on flange material was available for 228 (80.9%) pumps. Silicone was the most common material (*n*=205, 72.7%), followed by polypropylene (*n*=15, 5.3%), alone or combined with other materials (e.g., thermoplastic elastomer), and silicone combined with other materials (*n*=7, 2.5%). Fifty-four pumps (19.1%) did not specify the flange material.

Data on flange sizes were available for most pumps (*n*= 207, 73.4%). The reported flange sizes ranged from 15mm to 32mm. Size 24mm was the most frequently available flange size; included with 147 pumps (52.1%). Nineteen pumps (6.7%) marketed their flanges as “universal”, and two pumps provided generic sizes of “Small” and “Medium.” Only eight pumps (3.9%) provided five or more size options. As shown in **Figure 2a and Figure 2b** pump companies differed considerably in the range and combination of flange sizes offered. An evaluation of the co-occurrence network for flange sizes demonstrated that sizes 21 and 24 occurred most commonly together (weight = 60), followed by sizes 24, 19, and 21.

**Figure 2a:**
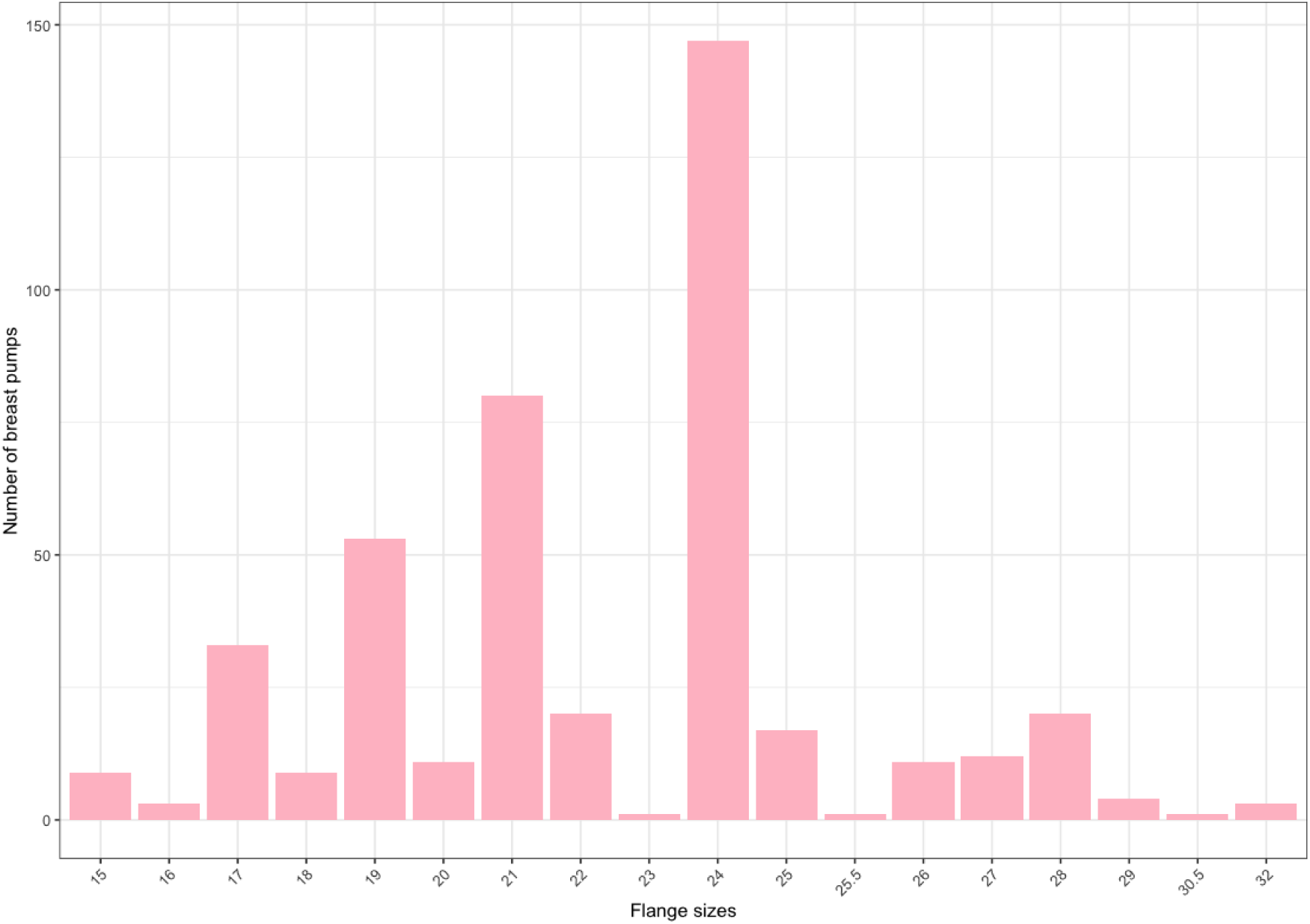
Frequency distribution of flanges sizes sold with commercial electric breast pumps^a^ ^a^ Only numerical flange sizes, ‘one size fits all’ were not included

**Figure 2b:**
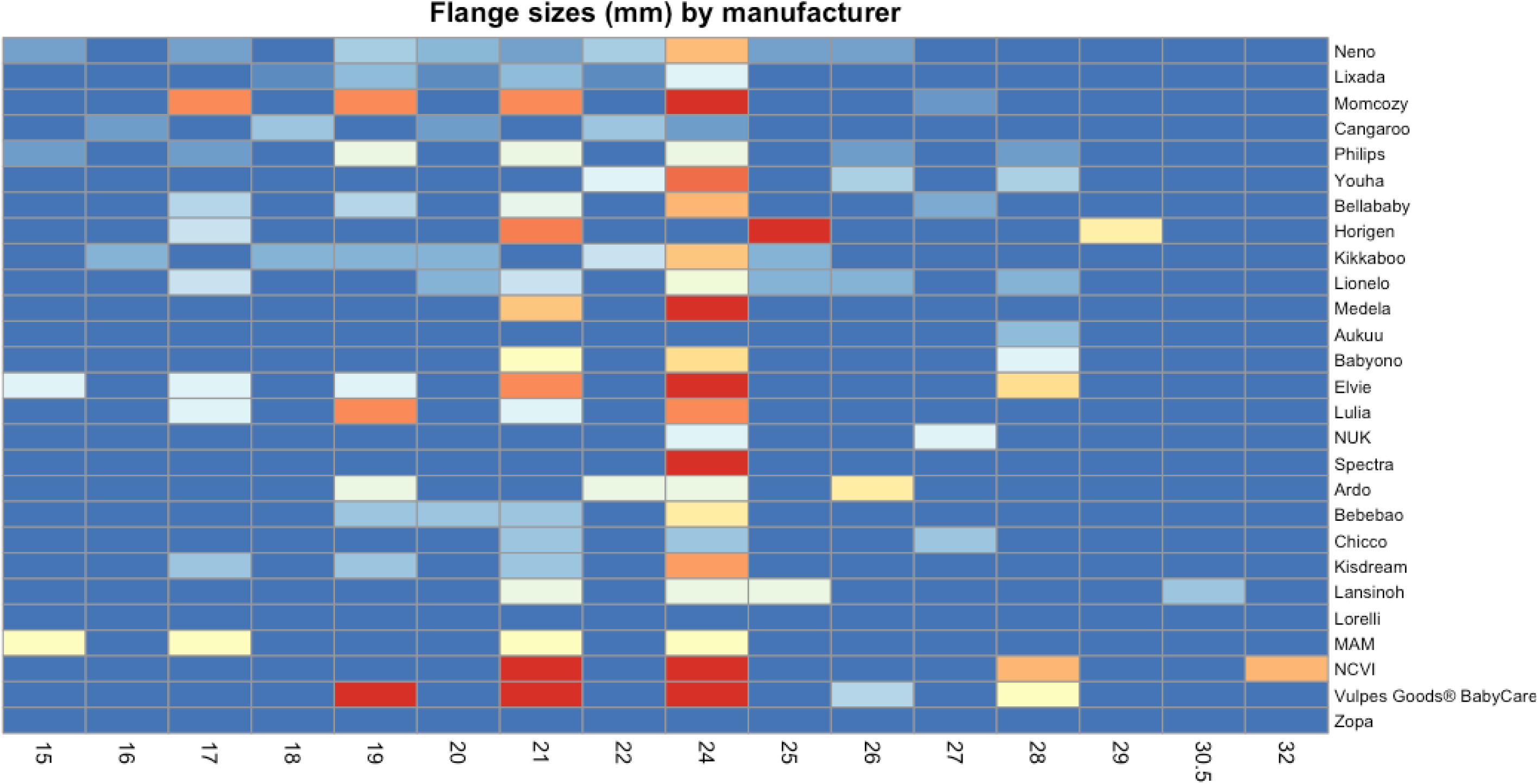
Heatmap of flange sizes offered by brands with four or more pumps

### 3.5 Retail cost of electric breast pumps

All 282 pumps had available price data with the price varying according to product configuration and discount rate. Prices varied widely (range: €19.99 - €679.57). Of the 282 pumps, 48 (17.0%) were discounted. The median list price was €76.35 (IQR €85.68), with a median discounted price of €72.73 (IQR €80.66, V =1172, p<0.001). The median absolute discount was €23.00 (IQR €37.98), corresponding to a median percentage discount of 17.2% (IQR 28.7%; range 2.0%-49.0%).

Prices significantly varied by certain pump characteristics (**see Figure 3: Boxplots of the distribution of prices by pump characteristics**). In addition to characteristics shown in the boxplots, battery capacity (ρ = 0.384, p < 0.001) and the number of flange sizes was positively correlated with the outcome (ρ = 0.254, p < 0.001). Pump volume demonstrated a weak negative correlation (ρ = -0.221, p < 0.001). No significant correlation was found for the number of intensity levels (ρ = 0.040, p = 0.516).

**Figure 3:**
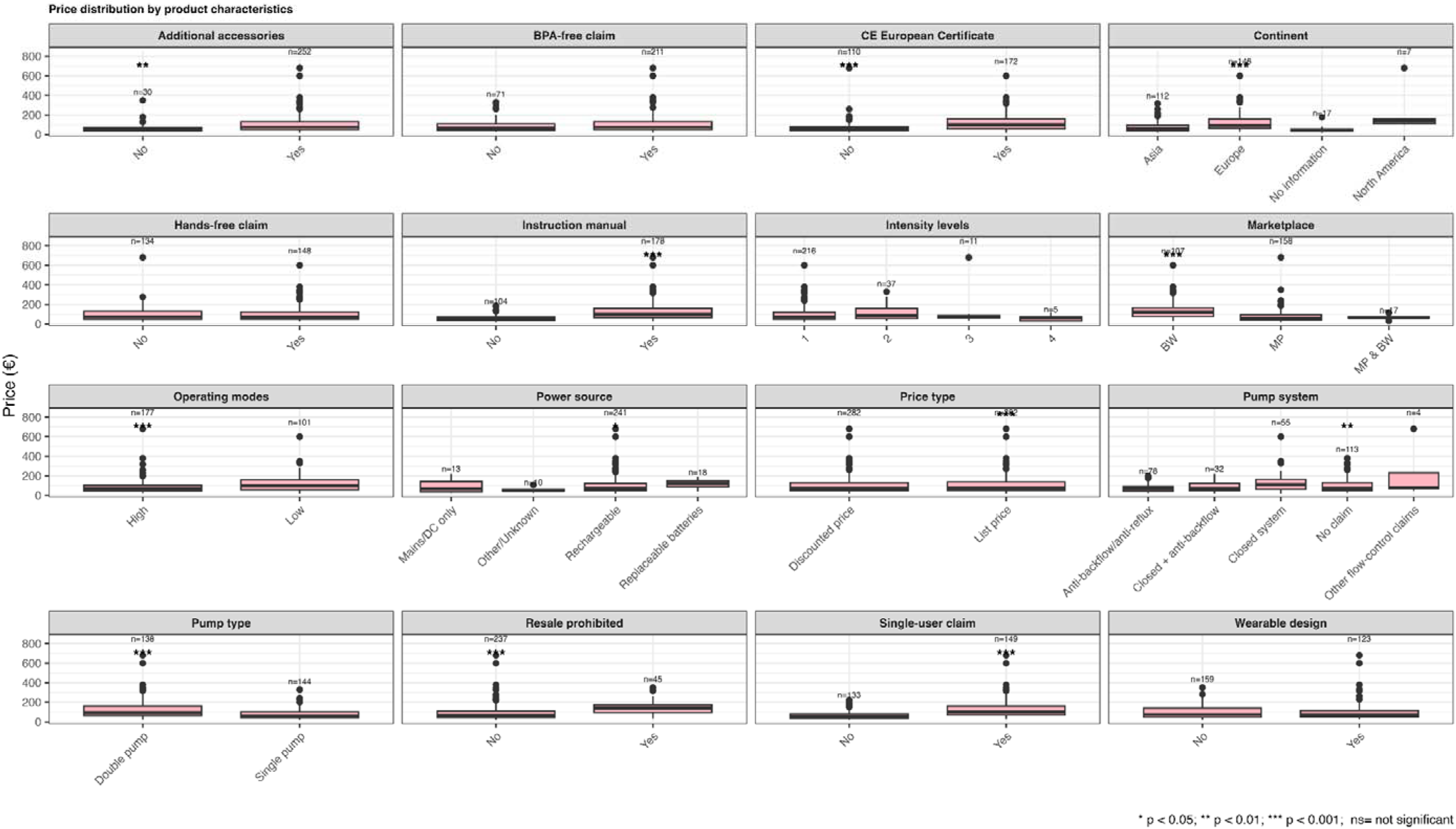
Boxplots of the distribution of prices by pump characteristics. Footnote: Define CE, * p<0.05, ** p<0.01, *** p <0.001, ns= not significant

Significant differences in price were observed between pumps with both single-user claims and an available instruction manual (χ² = 158.69, adjusted p < 0.001), instruction manual availability and continent (χ² = 85.08, adjusted p < 0.001), single-user claims and continent (χ² = 81.43, adjusted p < 0.001), and marketplace and instruction manual availability (χ² = 59.63, adjusted p < 0.001). Pumps with both a single-user claim and an instruction manual had the highest median price (€99.99, IQR €92.04), whereas those lacking both features had the lowest (€47.28, IQR €35.21). Pumps that were non-wearable, intended for single-person use, and did not advertise a hands-free function had a median price of €66.05 (IQR: €43.50–€109.60), whereas wearable double pumps that advertised hands-free operation had a higher median price of €99.99 (IQR: €71.49–€153.99). Overall, prices differed significantly across pump configurations (Kruskal–Wallis χ² = 39.5, df = 4, p < 0.001).

### 3.5 Predictors of price

Several predictors were associated with meaningful changes in price. Prices sourced exclusively through a digital marketplace were, on average, approximately 31% lower than those sourced through the brand website platform, and those available through both marketplaces and brand websites were about 26% lower. Double-sided pumps were 65% more expensive than single-sided models, while pumps with a “hands-free” claim were 31% more expensive than those without. Products with resale restrictions were 49% higher in price, whereas pumps with more pumping modes were 27% lower in price than those with less. Products from Asia were roughly 28% cheaper than those from Europe. Devices with four intensity levels were 43% less expensive than those with one.

The overall model contained a complete-case analysis of 282 products (*n*=200), was statistically significant and explained a substantial proportion of the variation in log price (adjusted R² = 0.65, (F (15, 184) = 25.26, p < 0.001). After variable selection, pumps that were resale prohibited, double-sided, and pumps from companies located in the European Union had significantly higher prices. These remained largely unchanged after multiple imputation (**see Table 3**, & supplementary Table 1).

## 4. Discussion

Breastfeeding and breast milk expression are not without costs—women who breastfeed bear both direct and indirect financial costs (6). This study found substantial costs associated with commercially available electric breast pumps. Direct costs associated with breast milk expression are a health equity issue, since these costs could be prohibitive to continuation of breastfeeding in mothers with socioeconomic constraints. Socioeconomic markers like low parental education have been shown to be associated with lower breastfeeding rates in cohorts in Germany (24, 25). Therefore, high direct costs for breast milk expression could potentially widen existing breastfeeding disparities and associated health outcomes (6, 18).

### Indirect costs associated with breastfeeding and breastmilk expression

Although paid parental leave is ensured by law in Germany, mothers with lower SES are disproportionately pressured to return to work earlier to lower-wage jobs (26). This phenomenon has been shown in other countries and is associated with premature breastfeeding cessation (27, 28). Acceleration in the separation of mothers and infants in the postpartum period heightens the uncompensated ‘opportunity costs’ of breastfeeding. Opportunity costs for breastfeeding mothers include the four to six hours per day dedicated to expressing breastmilk, time which might be otherwise spent on earning wages (6). In addition, self-employed women in Germany are not given paid time off. Whereas high-income professions tend to offer greater flexibility, such as greater autonomy and accommodations that support continued breastfeeding (6). For this reason, breastfeeding mothers in high-income professions might more easily balance breastfeeding with professional responsibilities (6). Comparatively, women working in low-wage positions might have limited workplace flexibility, unpredictable shifts, excessive workload, inadequate childcare facilities at work, and inadequate facilities for milk expression, making continued breastfeeding very challenging or impossible (27, 28). Consequently, structural socioeconomic inequalities are strengthened by unequal distribution of hidden labor costs which remain largely unrecognized and uncompensated by healthcare systems globally. Finally, the additional time required for feeding infants expressed breast milk (including storage, bottle-feeding, washing, and sterilization) compared to direct breastfeeding, should not be overlooked when calculating the full indirect cost burden associated with pumping.

### Coverage of direct costs for breast milk expression: benefits and remaining gaps

Reimbursement policies for direct costs associated with breast pumps in Germany differs from countries like the United States, where public health insurance covers the expenses of commercial electric breast pumps (29). Implementation of the Patient Protection and Affordable Care Act in the US requires new private insurance plans to provide breast pumps with no cost sharing since 2012 (30). This policy change showed trickle effects, demonstrating an uptick in the technological innovations in breast pumps (29), and an increase in the monthly rate of breast pump claims, which also translated into higher levels of breastfeeding for women with both private and public insurance (30). Breast pump use in the US context has in general been associated with a longer breastfeeding duration, even in marginalized women who are disproportionately affected by breastfeeding inequities (31). Despite healthcare coverage for commercial breast pump expenses, costs for breast milk expression for US families are not completely covered; parents report out-of-pocket expenditures for breastmilk storage bags, accessories and replacement parts (32).

### Direct costs related to breast pump characteristics and innovation

We found basic technical and location information for commercially available breast pumps was missing on digital marketplaces. Pumps that had more complete information were associated with higher prices. We suspect this is because regulatory compliance often requires investments in administrative processes, legal support, and customer service infrastructure. Products that are primarily produced in the EU showed a significant relationship with higher prices compared to those outside the EU. This supports the notion that availability of contact information and EU regulation may affect pump prices.

The heterogeneity in breast pump pricing is reflected in the very broad price range in our data set (range: €19.99 to €659.57). The observed variation in prices appears to reflect differences in underlying pump design, functionality, and intended use. Direct costs for the commercially available electric breast pumps surveyed varied significantly by source of information, manufacturer location, availability of accessories, double-sided pumping, power source, battery capacity, flange sizes, availability of instruction manuals, and “single user” claims, which all predicted significantly higher costs. Higher prices associated with some pump characteristics may be due to technological innovations or greater engineering complexity. Features such as double-pump capability, rechargeable batteries, hands-free operation, and milk collection systems require additional components and design considerations, which are likely to increase research and development and manufacturing costs and respective retail prices. Whether these characteristics translate into superior clinical or breastfeeding outcomes, however, is uncertain. One study of physicians using wearable pumps showed a longer breastfeeding duration, when compared to traditional pumps (33). There are comparatively few published pump efficacy studies encompassing a large sample of pumps, nor could we identify any health technology assessments in this area.

### Price and health equity considerations

These findings are relevant for mothers from lower SES who in the context of gaps in health insurance reimbursement in Germany might not be able to afford premium-priced pumps. Mothers with low SES are more likely to choose less expensive pumps because they are more affordable. However, lower-priced commercial breast pumps in our data set were statistically associated with less characteristics related to consumer accountability. For example, lower priced pumps were less likely to have a manufacturer website, manufacturer contact information, CE certificates, less likely to be manufactured in the EU, and in general were lesser-known brands with fewer pumps available on the market. These results have health equity implications, since mothers who purchase cheaper pumps in digital markets will receive less information about these pumps from manufacturers.

Price likely also indicates overall lower technological quality breast pumps; however, we are unable to draw strong conclusions related to the quality of commercially available breast pumps surveyed. This is because we did not directly measure technical performance indicators such as milk removal performance, accuracy of stated vacuum levels, battery capacity, or noise level. Therefore, we are unsure about the accuracy of reported information on digital marketplaces or how prices relate to the technical quality of commercially available pumps. Future studies should therefore explore whether cheaper pumps are of lower technical quality and whether they are associated with less maternal satisfaction or undesirable experiences with milk expression.

Our results demonstrated that double-sided commercially electric breast pumps were 65% more expensive than single-sided pumps. Although the direct costs associated with double-sided designs were higher, this additional upfront expenditure could translate to reduced indirect costs for mothers who pump their milk. Double-sided pumping has been shown to remove more milk and result in more milk ejections than pumping from each breast sequentially, thereby reducing the total pumping time (34). These indirect savings may include less time away from paid work or other responsibilities. Direct cost differences between single and double sided pumping raise important health equity considerations within the German healthcare system, which currently reimburses only one flange even with double sided electric breast pumps. This reimbursement structure effectively shifts indirect cost burden to families, which may disproportionately affect families with fewer financial resources.

There appears to be an increase in technological innovation in breast pumps, including wearable pump designs that allow for hands-free use, along with wireless and rechargeable functionality. We found that pumps with a “hands-free” claim were 31% more expensive than those without this feature or claim. While “hands-free” designs have higher direct cost for families, these designs may reduce certain indirect costs by allowing mothers to multitask while pumping, thereby lowering the opportunity cost of the time spent expressing milk, and allowing them to reach their breast milk feeding goals (33).

### Maternal experiences with commercial breast pumps

More studies on maternal experiences with pumping are needed, like comfort and degree of ease of use. Pumping does not only involve removing milk and feeding it to the baby, but managing all tasks around it, such as washing and sterilizing equipment, and managing milk storage (35). Surveys from higher income mothers have reported maternal needs related to pump portability, mobility, ease of use, ease of cleaning, fast and efficient milk extraction, comfort, discretion and low noise (32, 36).

### Food safety considerations

Personal use pumps hold the potential to cross-contaminate expressed breastmilk if mothers share or resell breast pumps intended for a single-user. However, second-hand online marketplaces list pre-owned personal-use breast pumps and related pump parts for resale. Breast pumps labeled as “not suitable for resale” therefore may reduce the risk of cross contamination. To our knowledge, the practice of resale of pre-used personal-use breast pumps and associated hygiene risks have not yet been empirically explored. In general, there is a paucity of studies on breast milk expression and pump hygiene in real-world settings (37). National guidance in Germany has been issued on the importance of proper hygiene when handling expressed milk and breast pumps at home and in clinic settings (38). However, hygiene aspects related to resale of personal use pumps and sharing pumps were not addressed. Therefore, our findings underscore gaps both in research and in policy guidance for consumer safety.

### Device safety

Many of the breast pumps surveyed did not have instructions for use available online. This is an important aspect for consumer safety, considering potential adverse events resulting from malfunction or improper use of breast pumps (39, 40). These findings correspond to other studies showing that oftentimes women are left to figure out pumping on their own, which shows women do not feel well-informed about how to use breast pumps to express milk (32, 41). Importantly, internet information on this topic may be inconsistent and not evidence-informed (42).

Improperly fitting flanges may cause or exacerbate nipple or breast pain, nipple trauma, and inadequate emptying of the breasts, which can, in turn, lead to infections or discontinuation of breastfeeding (15). Our findings demonstrate that commercially available breast pumps do not offer a range of flange sizes, with half of all pumps offering only a single flange size option. These findings highlight a critical gap in the availability of individualized flange sizes. Future research should evaluate how manufacturers, retailers, and health systems can improve access to appropriately fitting flanges to determine whether such efforts help reduce nipple trauma, pain, inadequate breast emptying, and early breastfeeding cessation.

### Limitations

Due to the cross-sectional study design, we were unable to capture volatility of price data over time. In addition, our study surveyed digital marketplaces but not brick and mortar stores. Therefore, we cannot make comparisons of costs of pumps online versus in stores.

## Conclusion

This study provides first insights into commercially available electric breast pumps in Germany and their associated prices. Commercial electric breast pumps sold on digital marketplaces have strikingly incomplete information that should therefore be more closely monitored by regulatory authorities. Accurate and complete information is of great importance for healthcare professionals and families and supports safe and appropriate use of medical devices (43, 44). Manufacturers should consider offering a broader range of breast-flange sizes to facilitate appropriate flange fit and provide information to consumers on appropriate flange fitting. We recommend health technology assessments and further studies on direct expenditures for infant feeding equipment in order to gain accurate estimations of the direct costs of infant feeding for families. Future health policy in this area should consider the substantial costs of breast pumps for families and the potential impact of current reimbursement policies on breastfeeding inequities in Germany.

## Declarations

No human subjects were involved in this research; therefore, ethical clearance was not required. JGA is a recipient of a scholarship from Medela, which was received after the work conducted at LMU Hospital for this study. All other authors declare no conflict of interest. Work on this study was funded by the Friedrich-Baur Foundation, the Bavarian Gender Equality Grant and the Ludwig-Maximilian-University of Munich (LMU) Hospital.

### Author Contributions

Conceptualization, methodology, funding: MT Writing: JGA, EO, FS, MZ, MT

Data collection and analysis

JGA, EO, MT Oversight and review: MT

### Acknowledgement of contributions

We acknowledge the contributions to piloting a data collection approach and support in drafting the study protocol by Nina Berg. We acknowledge Theresa Stegeman for her help with data collection.

## Acknowledgement of AI

Wording refinement was supported by an artificial intelligence tool (Microsoft Copilot). The authors reviewed and approved all content, and all interpretations and conclusions were made by the research team.

## Data Availability Statement

The data used in this research will be available from the authors upon reasonable request.

