## Supplementary Figure 1 for "Determinants of prices of commercial electric breast pumps sold in Germany: a cross-sectional analysis"

**Supplementary Figure 1: Overview of Missing and Observed Data for Commercial Pumps on German Digital Marketplaces**

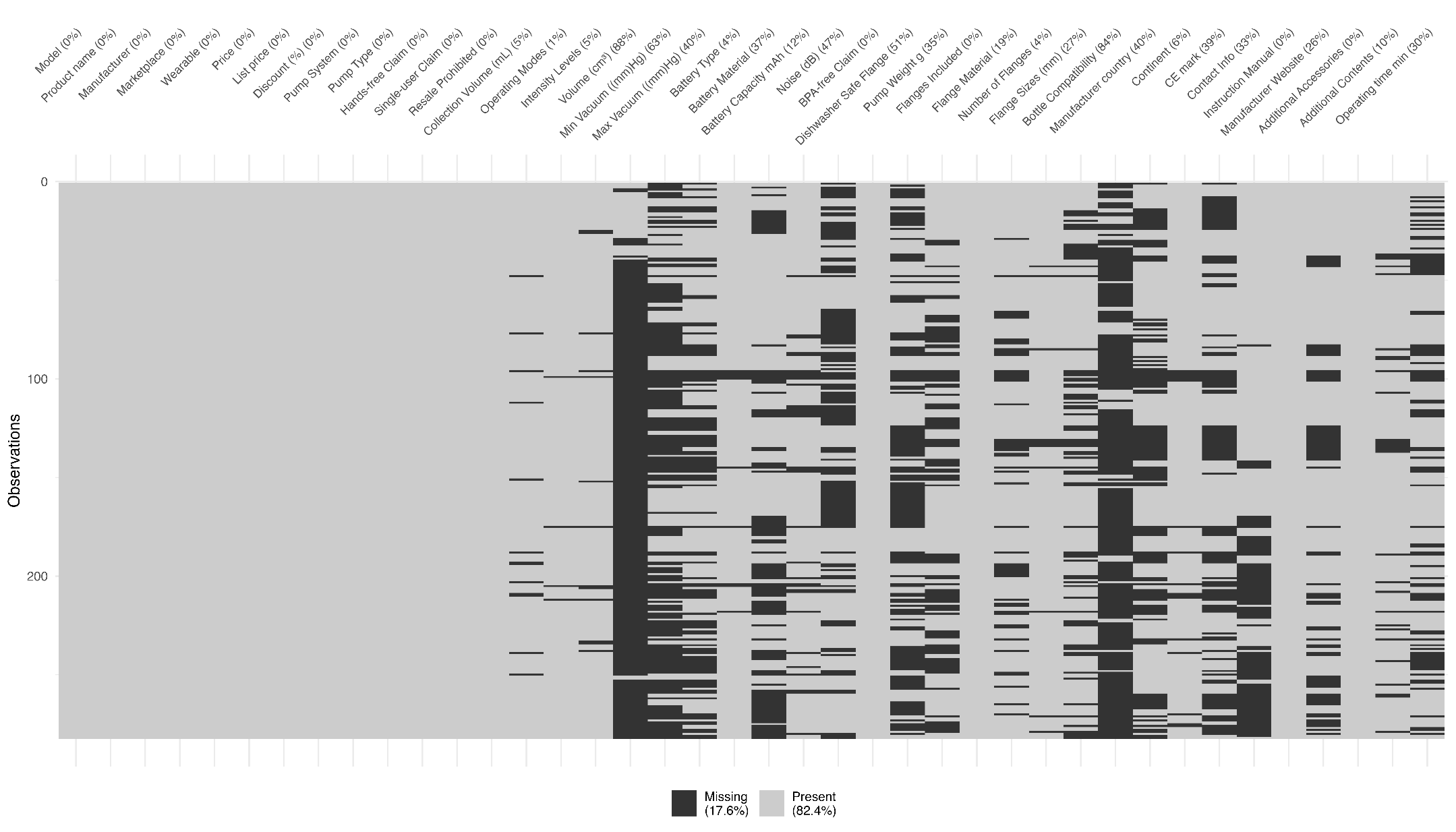
