## Supplementary Figure 2 for "Determinants of prices of commercial electric breast pumps sold in Germany: a cross-sectional analysis"

**Supplementary Figure 2: Flowchart of selection process for commercial electric breast pumps on digital marketplaces**

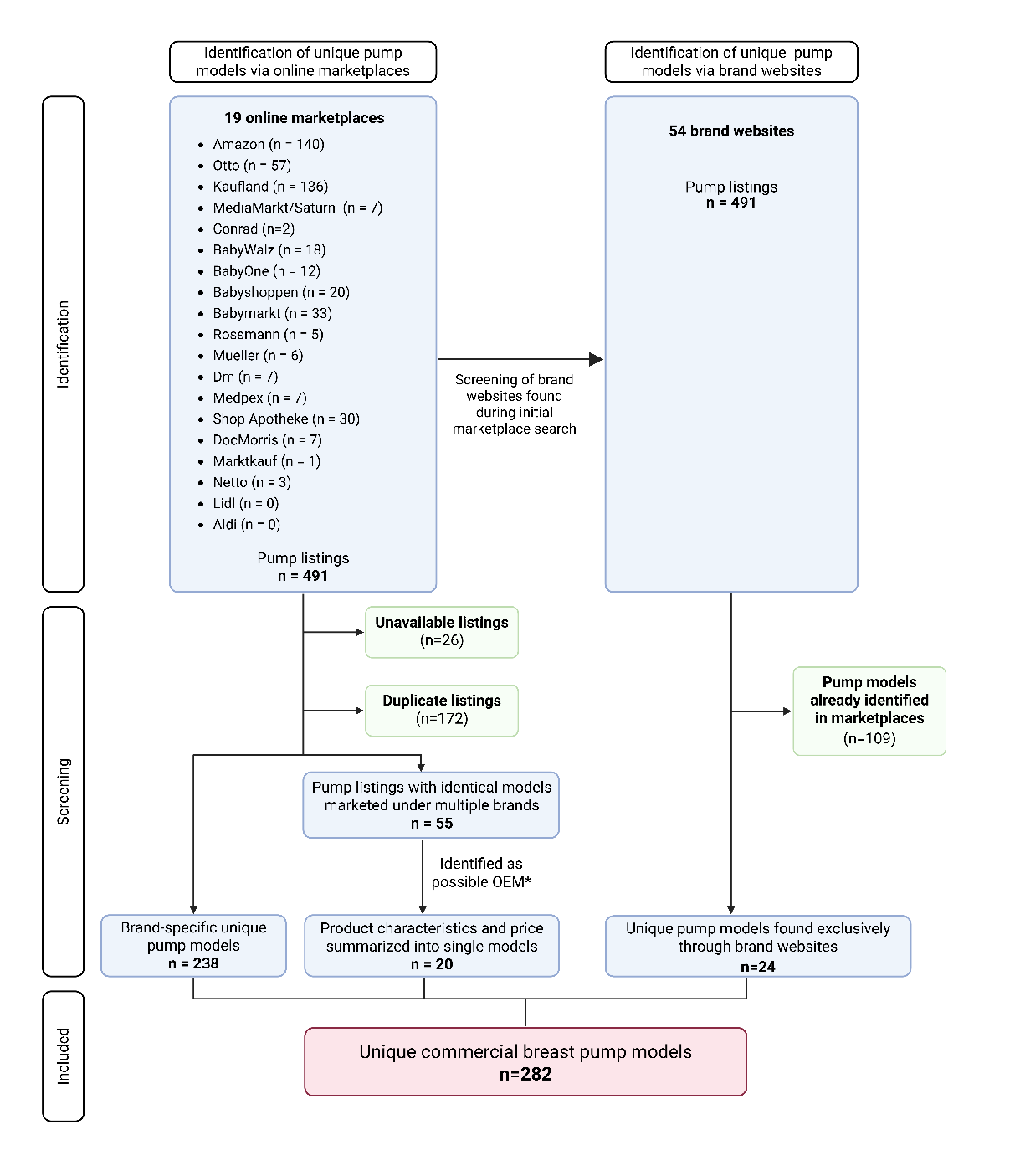

*OEM: Original Equipment Manufacturer
