## Supplementary Figure 3 for "Determinants of prices of commercial electric breast pumps sold in Germany: a cross-sectional analysis"

**Supplementary Table 1: Breast pump brand headquarters and manufacturing country information**

| **No** | **Brand** | **Brand Headquarters Location** | **Manufacturing country** | **Website** |
| --- | --- | --- | --- | --- |
| 1 | Aiorber | no information | no information | no website |
| 2 | Andoer | no information | no information | no website |
| 3 | Aoucheni | no information | no information | no website |
| 4 | Ardo | Ardo medical AG  Gewerbestrasse 19  6314 Unterägeri,  Switzerland | Switzerland | <https://shop.ardo.de/>  https://ardomedical.com/ |
| 5 | Aukuu | no information | no information | no website |
| 6 | Babyhit Basics | Kobylinska 40B  63-760 Zduny,  Poland | no information | https://babyhit.de/ |
| 7 | Babyono | 13 Kowalewicka Street  60-002 Poznań  Poland | no information | https://babyono.com/en |
| 8 | Banlvs | no information | no information | no website |
| 9 | Beaba | 37 rue de Liège, 75008 Paris, France | China | https://www.beaba.com/de-de |
| 10 | Bebebao | no information | China | no website |
| 11 | Bebeconfort (Brand owned by Dorel Juvenile) | Korendijk 5,  5704 RD Helmond,  The Netherlands  (EU head office of Dorel Juvenile) | France | https://www.bebeconfort.co.de |
| 12 | Bellababy | Ningbo Huiyoo Baby Products Co. Ltd. 523# Cizhang Road, Zhangqi Industrial Area,  Cixi, 315313,  Zhejiang,  China | China | https://bellababy-med.com/ |
| 13 | Beurer | Beurer GmbH Söflinger Strasse 218  89077 Ulm  Germany | China | https://www.beurer.com/de/ |
| 14 | Bimirth | no information | no information | no website |
| 15 | Bioby | Hong Kong Ting Hui Limited  21/F, Hing Lung Commercial Building, 68-74 Bonham Strand, Sheung Wan,  Hong Kong | China | no website |
| 16 | Bocioland  (Owned by Top Sale Sp. z o. o.) | Grzybowska 87, 00-844 Warsaw,  Poland | no information | https://bocioland.pl |
| 17 | Bremen | 706C, 7/F, Hao Sheng Business Center, Dongbin  Road, Nanshan Street, Nanshan District  518000 Shenzhen  China | China | no website |
| 18 | Ca-mi | Via Ugo La Malfa 13 – Frazione Pilastro  43013 Langhirano  Parma  Italy | no information | https://www.ca-mi.eu |
| 19 | Cangaroo | Moni Trade Ltd.  1 Dolo Street  1298 Sofia  Bulgaria | Bulgaria | <https://cangaroo-bg.com> // <https://moni.bg/> |
| 20 | Canpol Babies | 430 Puławska street,  02-884 Warsaw,  Poland | Poland | https://canpolbabies.com/gb_EN/products/breastfeeding/breast-pumps |
| 21 | Chicco | Artsana S.p.A  via Saldarini Catelli, 1 - 22070 Grandate (Como) Italy. | China | https://www.chicco.de/ |
| 22 | Cool-i | No. 23, Nanfeng, Dajiang Road, Leliu Town  Shunde District,  Foshan City,  Guangdong Province,  China | China | no website |
| 23 | Decdeal | no information | no information | no website |
| 24 | Dopwii | no information | no information | no website |
| 25 | Dr. Brown’s | Dr. Brown's Company Inc. 4433 Fyler Avenue St. Louis, Missouri 63116  USA | China | https://drbrownsbaby.com/ |
| 26 | Dreamhigh | Room B1414,  Gediao Building, Guangkaisima Road,  Nankai District,  Tianjin,  China | China | no website |
| 27 | DTC Gmbh | no information | no information | no website |
| 28 | Easybaby | Handelsweg 8a  3881 LS / Putten  The Netherlands | China | https://www.easybaby-shop.de |
| 29 | Elvie | Chiaro Technology Limited  63-66 Hatton Garden EC1N 8LE London United Kingdom | Singapore, China | https://www.elvie.com/de-de |
| 30 | Esperanza | 05-850 OŻARÓW MAZOWIECKI, ul. Poznańska 129/133  Poland | China | https://esperanza.pl/index.php |
| 31 | eSynic | RM18G 27/F Ho King Comm CTR,  2-16 FAYUEN St Mongkok,  Kl Hong Kong 999077,  China | Hong Kong | https://esynic.com/ |
| 32 | Eta | Křižíkova 148/34  Praha 8 - Karlín, 186 00  Czech Republic | no information | https://eta-hausgeraete.de/ |
| 33 | Extralink | ul. 1 maja 23, 42-217, Częstochowa,  Poland | no information | https://www.extralink24.com/ |
| 34 | Fiqops | no information | no information | no website |
| 35 | Flaem | Via Colli Storici nr. 221/223/225 25015 Martino della Battaglia - Brescia  Italy | Italy | https://www.flaem.eu/en/ |
| 36 | Funmi | 50 Ferny Croft  Haywards Heath  West Sussex  RH16 4UP  United Kingdom | China | https://shopfunmi.com/ |
| 37 | Grottes | no information | no information | no website |
| 38 | Hanience | no information | no information | no website |
| 39 | Heyvalue | no information | no information | https://www.heyvaluestore.com/ |
| 40 | Horigen | No. 18,  Pingnan Industrial Zone,  Mianbei Street,  Chaoyang District,  Shantou, Guangdong  515100,  China | China | https://horigen.net/ |
| 41 | HyAdierTech | Chengdu Lidao  [Floor 6, Unit 3, Building 7, No.399, West Section, Fucheng Avenue,](https://maps.google.com/?q=No.607,%20Floor%206,%20Unit%203,%20Building%207,%20No.399,%20West%20Section,%20Fucheng%20Avenue,%20Chengdu%20High-te%20ch%20Zone,%20China%20Pilot%20Free%20Trade%20Zone%20Chengdu,%20Sichuan,%20610014%20China)  [Chengdu High-tech Zone, Chengdu, Sichuan,](https://maps.google.com/?q=No.607,%20Floor%206,%20Unit%203,%20Building%207,%20No.399,%20West%20Section,%20Fucheng%20Avenue,%20Chengdu%20High-te%20ch%20Zone,%20China%20Pilot%20Free%20Trade%20Zone%20Chengdu,%20Sichuan,%20610014%20China)  [610014](https://maps.google.com/?q=No.607,%20Floor%206,%20Unit%203,%20Building%207,%20No.399,%20West%20Section,%20Fucheng%20Avenue,%20Chengdu%20High-te%20ch%20Zone,%20China%20Pilot%20Free%20Trade%20Zone%20Chengdu,%20Sichuan,%20610014%20China)  [Chin](https://maps.google.com/?q=No.607,%20Floor%206,%20Unit%203,%20Building%207,%20No.399,%20West%20Section,%20Fucheng%20Avenue,%20Chengdu%20High-te%20ch%20Zone,%20China%20Pilot%20Free%20Trade%20Zone%20Chengdu,%20Sichuan,%20610014%20China)a | China | no website |
| 42 | Ikare | Room 803,  Chevalier House 45-51  Chatham Road South  Tsim Sha Tsui, Kowloon,  HongKong, 200030,  China | China | https://www.ikare.vip/ |
| 43 | Jiubiz | no information | no information | no website |
| 44 | Jopassy | no information | no information | no website |
| 45 | Kikkaboo | 4000 Plovdiv,  str. Vasil Levski 121  Bulgaria | China | https://kikkaboo.com/ |
| 46 | Kisdmxf | no information | no information | no website |
| 47 | Kisdream | 12 Orchard Street,  Bristol,  United Kingdom | no information | https://kisdream.com/ |
| 48 | Kissbobo | Jun Yang Technology Co., Ltd. - Flat/Rm A, 12/F, KIU FU Commercial Building, 300 Lockhart Road, Wan Chai, Hong Kong | China | https://kissbo.com/de |
| 49 | Lansinoh | 333 North Fairfax Street, Suite 400, Alexandria, VA 22314, USA | China/Turkey | https://lansinoh.de/ |
| 50 | Letgospt | no information | no information | no website |
| 51 | Lionelo | ul. Adama Kręglewskiego 1, 61-248 Poznań,  Poland | China | https://lionelo.de |
| 52 | Lixada | Guangdong Province, China | China | no website |
| 53 | Lola & Lykke | Oy Konepajankuja 1  00510 Helsinki  Finland | no information | https://lolalykke.de/ |
| 54 | Lorelli | Didis Ltd., 6 Trakia-Iztok Street, Shumen,  Bulgaria | China | https://lorelli.eu/en-bg/ |
| 55 | Lovi | ul. Graniczna 4, Słubica B  96-321 Żabia Wola  Poland | no information | https://lovibaby.com/gb_EN |
| 56 | Lulia | Shenzhen Meide Hongda Technology Co.,Ltd  Room 1308, Building A, Yonghuayuan No. 6,  Baotian 2nd Road,  Chentian Community,  Xixiang Street,  Bao'an District,  Shenzhen City,  Guangdong Province,  China | China | no website |
| 57 | Magcubic | Hunan Aibeisite Technology Co., Ltd.5F, Building 2, Runfeng Zhigu, No. 3 Sima Gongye 1st Road,  Changping Town, Dongguan, Guangdong, China | China | no website |
| 58 | MAM | Lorenz-Mandl-Gasse 50  1160 Vienna,  Austria | Hungary, Thailand | https://www.mambaby.com/de/ |
| 59 | Mamajoo | Mimar Sinan Mahallesi, Özgürlük Caddesi No: 77/1, 34935 Sultanbeyli, Istanbul,  Turkey | Turkey | https://www.mamajoo.com/ |
| 60 | Medela | Medela AG  Lättichstrasse 4b  6341 Baar  Switzerland | China, USA, Switzerland | https://www.medela.com/de-de/stillen-und-abpumpen |
| 61 | Microlife | Microlife AG Swiss Corporation  Espenstrasse 139, CH-9443 Widnau/ Switzerland | China, Taiwan, Switzerland | https://www.microlife.com/ |
| 62 | Missaa | Shenzen TPH Technology Co., Ltd.  East side of 5th floor, Building 29, Phase 2, Lianchuang Science and Technology Park,  Bulan Road, Nanwan Street, Longgang District,  Shenzhen  China | China | no website |
| 63 | Momcozy | 1624 Market St,  Ste 226, #30327,  Denver,  CO 80202,  USA | China | https://de.momcozy.com/ |
| 64 | MomMed | Room 803 45-51 Chatham Road South, Tsim Sha Tsui, Kowloon,  Hongkong | no information | https://mommed.com/en-eu |
| 65 | Mopuea | no information | no information | no website |
| 66 | Mumgaroo | no information | China | no website |
| 67 | NCVI | Jiangsu Xinbei Electrical Appliances Co., Ltd.  No.115,Xinjin Road, Meicun,  Xinwu District,Wuxi City, Jiangsu Province  China | China | https://www.ncvi.vip/ |
| 68 | Neno | 31-752 5b Ujastek street, Kraków,  Poland | China | https://neno.baby/ |
| 69 | Neovsuny | no information | no information | no website |
| 70 | Novzep | no information | no information | no website |
| 71 | NUK | MAPA GmbH  Industriestraße 21-25  D 27404 Zeven  Germany | China, Germany | https://www.nuk.de/ |
| 72 | Opnmina | no information | no information | no website |
| 73 | Oyajia | no information | no information | no website |
| 74 | Perifit | 128, Rue de la Boétie  75008 Paris,  France | no information | https://de.perifit.co/ |
| 75 | Philips Avent | Amstelplein 2, 1096 BC Amsterdam,  The Netherlands  (Originally a UK brand) | China, Hungary | https://www.philips.de/ |
| 76 | PiAEK | Ningbo Qinmeng Maternal and Infant Products CO., LTD  No.202 Guyao Road, Guyaopu Village, Zhangqi Town, Cixi City, Zhejiang China. | China | no website |
| 77 | Sejoy | Sejoy / Joytech (Sejoy Biomedical / Joytech Healthcare  No. 365 Wuzhou Road  (Area C, Building 2),  Yuhang Economic Development Zone, Hangzhou City,  Zhejiang Province, 311100,  China | China | https://www.sejoygroup.com/de/ |
| 78 | Severno | no information | no information | no website |
| 79 | Spectra | Uzinmedicare / Uzinmedical origins  54, Dongtansandan 5-gil, Hwaseong-si, Gyeonggi-do, 18487  South Korea | South Korea | https://www.spectra-milchpumpen.de/? |
| 80 | Sross | no information | China | no website |
| 81 | Startech | no information | no information | no website |
| 82 | Suavinex | Laboratorios Suavinex  Calle del Marco, parcela R88 (Polígono Las Atalayas) – 03114 C/ Torreblanca, 10, 03006 Alicante,  Spain | China | https://www.suavinex.com/ |
| 83 | Swanew | no information | no information | no website |
| 84 | Tianpeng | Shenzhen City Tianpeng Technology Development Co., Ltd.  Room 9B, Building 8, Shidaijingyuan, Baishixia Community, Fuyong Street, Baoan District, Shenzhen City, Guangdong Province,  China | China | no website |
| 85 | TolleTour | no information | no information | no website |
| 86 | Tommee Tippee | Mayborn UK Limited  Mayborn House,  Balliol Business Park,  Benton Lane,  Newcastle Upon Tyne,  NE12 8EW,  United Kingdom | China | https://www.tommeetippee.com/en-gb/ |
| 87 | Tovvild | no information | no information | no website |
| 88 | Truelife | elem6 s.r.o., Braškovská 308/15, 161 00 Prague 6,  Czech Republic | no information | https://truelife.eu/en |
| 89 | Ukuu | no information | China | no website |
| 90 | Uleway | no information | China | no website |
| 91 | Urmywo | no information | China | no website |
| 92 | Uwot | no information | no information | no website |
| 93 | Vayotoy | [Xianxia Medical Technology Co., Ltd.](https://www.google.com/search?sca_esv=5b71cfa89013eb9f&cs=0&sxsrf=AE3TifOtncLaRh0nnVF6vWthukjQ5WH3VA%3A1757937638619&q=Xianxia+Medical+Technology+Co.%2C+Ltd.&sa=X&ved=2ahUKEwij4dqD3NqPAxWoFBAIHeEVBM4QxccNegQIAxAB&mstk=AUtExfDrgWbCwWRlh63jCTuTFdVloTSv1WmnzdG6vFXiH_EW3wFkYrVPLR0-zxN6sdCQ-EtR2V890q8aYJqwYxxfM80mY39Z-aWAvN0PNWvPsS-kwqTt48S3S6edIIwLj7r2h-MEPsmSqiDjxmADWMNz0HISOUAPzFzHUrESrmluuD33IXVc_RyFAKNRuEEKO3aAC7urNWEotlZy_EHleYLZDSoAOkia-V_NXT-LqfNaSYGHn_bV-3S7371kg6KtFrvi_3uV8OvEDTqjqQYrJ_huZ-pJ&csui=3),  6501 CTF Finance Centre, No.6 Zhujiang Road East, Tianhe District, Guangzhou 510623, China | China | no website |
| 94 | Vevor | Baoshanqu Shuangchenglu 803long 11hao 1602A-1609shi Shanghai,  China | China | https://www.vevor.de/ |
| 95 | Vorsprung | DropNet LTD 2,  Neville Place Company number 13222370.  London,  N22 8HX  United Kingdom | China | https://vorsprung.com/en-eu |
| 96 | Vulpes Goods Baby Care | Veilingweg 44,  3981PC Bunnik,  The Netherlands | no information | https://www.vulpesgoods.de/ |
| 97 | Welikera | Shenzhenshi kehuishiyan keji youxiangongsi. Baoanqu Xixiangjiedao Xixiangdadao Baoyuanhuafengzongbu Jingjidasha B zuo 531shi. Shenzhen Guangdong,  China | China | no website |
| 98 | Wilgoon | no information | no information | no website |
| 99 | Willow | Willow Innovations, Inc.  1975 W. El Camino Real, Suite 306, Mountain View, CA 94040  USA | Taiwan | https://onewillow.com/ |
| 100 | Xocome | no information | China | no website |
| 101 | Yabtf | no information | China | no website |
| 102 | Yagosodee | no information | China | no website |
| 103 | Yakimz | no information | no information | no website |
| 104 | Youha | No. 567, Xintang Road, Fuhai Town, Cixi City, Ningbo City, Zhejiang Province, China | China | https://www.youha.com/about-us.html |
| 105 | Yozhiqu | no information | no information | no website |
| 106 | Zggzerg | no information | no information | no website |
| 107 | Zimtky | no information | no information | no website |
| 108 | Zopa | ZOPA design s.r.o.  Záhřebská 562/41  120 00 Prague  Czech Republic | no information | https://zopadesign.com/en/ |
| Total: | 108 | Total brands with address information: 66 | Total brands with at least one country of manufacturing: 61 | Total brands with website: 54 |
